# Presence of additional *P. vivax* malaria in Duffy negative individuals from Southwestern Nigeria

**DOI:** 10.1101/19009837

**Authors:** Mary Aigbiremo Oboh, Upasana Shyamsunder Singh, Daouda Nidaye, Aida S. Badiane, Anwar Ali, Praveen Kumar Bharti, Aparup Das

**Affiliations:** Parasitology and Mycology Laboratory, Université Cheikh Anta Diop, Dakar, Senegal; Genomic Epidemiology Laboratory, Division of Vector Borne Diseases, ICMR-National Institute of Research in Tribal Health, Jabalpur, India; School of Earth and Environmental Sciences, University of Manchester, United Kingdom

## Abstract

Malaria in sub-Saharan Africa (sSA) is thought to be hugely caused by *Plasmodium falciparum* and very infrequently by *P. ovale, P. malariae*, with *P. vivax* not even being considered to be of any significant role. However, with the availability of very sensitive diagnostic tool, it has become more clear that, the percentage of non-falciparum malaria in this sub-region has been underestimated. *P. vivax* was historically thought to be absent in sSA due to the high prevalence of the Duffy null antigen in individuals residing here. Nevertheless, recent studies reporting the detection of vivax malaria in Duffy-negative individuals challenges this notion. Following our earlier report of *P. vivax* in Duffy-negative individuals, we have re-assessed all previous samples following the classical PCR method and sequencing to confirm both single/mixed infections as well as the Duffy status of the individuals.

Interestingly, fifteen additional *Plasmodium* infections were detected, representing 5.9% in prevalence from our earlier work. In addition, *P. vivax* represents 26.7% (4/15) of the new isolates collected in Nigeria. Sequencing results confirmed, all vivax isolates as truly vivax malaria and their Duffy status to be that of the Duffy-negative genotype. The identification of more vivax isolates among these Duffy-negative individuals from Nigeria, substantiate the expanding body of evidence of the ability of *P. vivax* to infect RBCs that do not express the DARC gene. Hence, such geno-epidemiological study should be conducted at the national level in order to evaluate the actual burden of *P. vivax* in the country.

## INTRODUCTION

Malaria is a critical infectious disease of public health importance that provokes considerable mortality in all endemic countries. The tremendous gains seen in cases and mortality reduction is as a result of deliberate intervention strategies (1). However, the observed benefits has seen a plateau in the last two years especially in Africa where the greatest burden of disease is mostly impacted. In sub-Saharan Africa (sSA), majority (99%) of the infections is thought to be due to *Plasmodium falciparum* and very infrequently by *P. ovale, P. malariae*, with *P. vivax* not even being considered in the picture as one of the players (1). With the availability of tools that are more sensitive, the detection of non-falciparum and even vivax human malaria parasites has gained more attention in sSA (2–6).

Historically, *P. vivax* prevails in Asia, (7,8), South America (9,10) and has some scanty presence in the Horns of Africa such as in Djibouti (11), Eritrea (12), Somalia (13,14), Ethiopia (15–18) and Sudan (19,20). Thus *P. vivax* has a much wider geographical distribution unlike falciparum malaria which in a more specific term, can be said to have a much focal distribution in Africa.

Hence, the former notion is that, *P. vivax* originates from Asia and South America and gradually finds its way into Africa through the trade-route corridor. However, there are some current evidences supporting the hypothesis that *P. vivax* could have originally evolved from a vivax-like strain detected in non-human primates in Africa (21,22) and from there, dispersed to other continents during the period of human migration. Although, both hypotheses (whether from Africa to Asia or Asia to Africa) require further validation. However, it seems likely that there might be an interplay of both hypothesis, in which case, simultaneous occurrence takes place and balancing selection of the Duffy negative gene in sSA might have resulted in the absence of vivax malaria in the region. Nevertheless, later re-introduced when individuals expressing the Duffy gene travels between continents and countries.

The Duffy (gp-*FY;* CD234) gene is the fourth red blood cell (RBC) gene after thalassemia, sickle cell anaemia and glucose-6-phosphate dehydrogenase (G6PD) associated with resistance to *Plasmodium* species (23) with particular protection against vivax malaria. Also known as the Duffy antigen receptor for chemokines (DARC), is a variable receptor usually expressed on the surface of the red blood cell (RBC) and employed by *P. vivax* merozoites in gaining access in the RBCs and establishing its erythrocytic infection (24). The DARC, located on chromosome 1 has two exons, and a single nucleotide substitution from a thymine (T) to a cytosine (C), upstream in the promoter region nullifies the expression of this gene on the RBCs, resulting in the FYO* allele (25). This FYO* null allele predominates amongst sSA inhabitants as with African-Americans but has a very sparse representation in individuals of other ancestry (26). Thus, the FYO* null allele has been postulated to confer protection against *P. vivax* infection in this sub-region. Nevertheless, 11 countries in this region (Oboh et al unpublished data) have reported the occurrence of *P. vivax*, making it more real that vivax malaria might be gradually finding its way into sSA, thus it can be postulated that hidden transmission is occurring in this region. In some of these studies, such as in those conducted in Angola, Cameroon, Kenya, Madagascar, Mali and Mauritania, the Duffy status of the infected individuals was characterized and they were found to be mostly Duffy negative (3,4,27–30). In others, however, the investigators just stopped at identification of *P. vivax* without stating the Duffy status of the infected individuals (5,31– 33). Interestingly, all studies were carried out amongst indigenous individuals with little or no travel history to vivax endemic areas, ruling out the possibility of imported infection.

In Nigeria, *P. falciparum* is responsible for > 95% of malaria infection, with *P. malariae* and *P. ovale* contributing a meagre < 5% of infection (1,34). Data implicating *P. vivax* infection in Nigeria includes its detection in a visiting pregnant female (35), two microscopical cases (36,37), both of which were not confirmed by any molecular technique and our previous study (6) which detected five Duffy negative individuals to be infected with *P. vivax* isolates and were subsequently confirmed by capillary sequencing.

Thus, as a follow-up to our previous study, we have re-evaluated all past samples using the classical PCR method, to confirm additional *P. vivax* isolates (both single and mixed infection) by sequencing as well as determined the Duffy status of the individuals. The importance of such genomic epidemiological studies cannot be undermined in this era of malaria elimination, as attention also needs to be given to non-falciparum infection, if the ambitious albeit, achievable 2030 elimination goal is to be reached.

## Materials and Methods

Blood samples were collected from symptomatic patients attending four different hospitals in Lagos (Gbagada, Ikorodu, Akodo and Ikate) and two in Edo (Central and Stella) states respectively, and quickly subjected to malaria rapid diagnostic test kit, employing the manufacturer’s instruction (Pf-HRPII-Care Start®, Access Bio Inc, Batch number M014L04-M014M10) followed by microscopy (results for both not shown here).Two dried blood-spots per patient (436 in total), irrespective of their status (positive or negative by any of the technique above) were made on Whatmann® (GE Healthcare, Life Sciences) filter paper and brought to the ICMR-National Institute of Research in Tribal Health (ICMR-NIRTH), Jabalpur, India, after obtaining appropriate ethical clearance (IRB/16/347) from the Nigerian Institute of Medical research.

Employing the Qiagen®kit (the QIAamp DNA Blood Mini Kit; Hilden, Germany), genomic DNA was re-isolated from all 436 samples and subsequently subjected to nested PCR diagnostic protocol targeting the 18S rRNA to identify all four *Plasmodium* infecting species using primer pairs as designed earlier (Snounou and Singh, 2002) (Supplementary Table 1).For each PCR run, a negative control (nuclease free water) and positive controls (sequenced confirmed *Plasmodium* species-for all four species) were added. Additionally, a part of the Duffy gene promoter region (for isolates that are *P. vivax* positive) was PCR amplified and sequenced in order to determine their Duffy status using protocols and primer sequence detailed in Supplementary Table 2.

Representative isolates of *Plasmodium* species (*P. falciparum, P. vivax, P. malariae* and *P. ovale*) were purified (using Fast^AP^ alkaline phosphatase and exonuclease I) and processed for sequencing by Sanger method (an in-house facility of ICMR-NIRTH, Jabalpur) in both direction (2X).Sequencing was performed on the purified PCR products in a volume of 10µl with 0.5µl of Terminator ready reaction mix (TRR), 1.6 pmol of gene specific primer and 5X reaction buffer with a cycling condition of 96°C denaturation for 10secs (25 cycles), annealing at 50°C for 5sec and an extension of 60°C for four minutes. Base calling of nucleotide and chromatogram visualization was achieved with the use of the sequence analysis software accompanying the DNA analyzer, while sequence alignment was carried out using BioEdit sequence alignment editor v.7.0.5.3. Contiguous sequence were aligned with their respective reference strains (*P. vivax* -SAL-1 accession number U03079.1; *P. falciparum*-3D7 accession umber XR_002273095.1; *P. ovale*-accession number L48987.1; *P. malariae*-accession number NG_011626.30 and, the Duffy gene; accession number NG_011626.30).

## Results and Discussions

*Plasmodium* genomic DNA was amplifiable in 62.2% (271/436) of the isolates, representing a 5.9% (15) increase in the PCR amplifiable products from previous study (58.7%; 256/436) (6), with *P. falciparum* being the highest detectable species (40%: 6/15), interestingly followed by *P. vivax* (26.7%:4/15), *P. falciparum*/*P. ovale* mixed infection (26.7%:4/15) and only one 6.6%)*P. malariae* was additionally identified. In attempts to rule out false positive, the gDNA of the newly identified *P. vivax* isolates were re-extracted and amplified twice following same protocol earlier used (see supplementary Table 1). Gel documentation of all newly identified isolates is presented in Figure 1.

**Figure 1:**
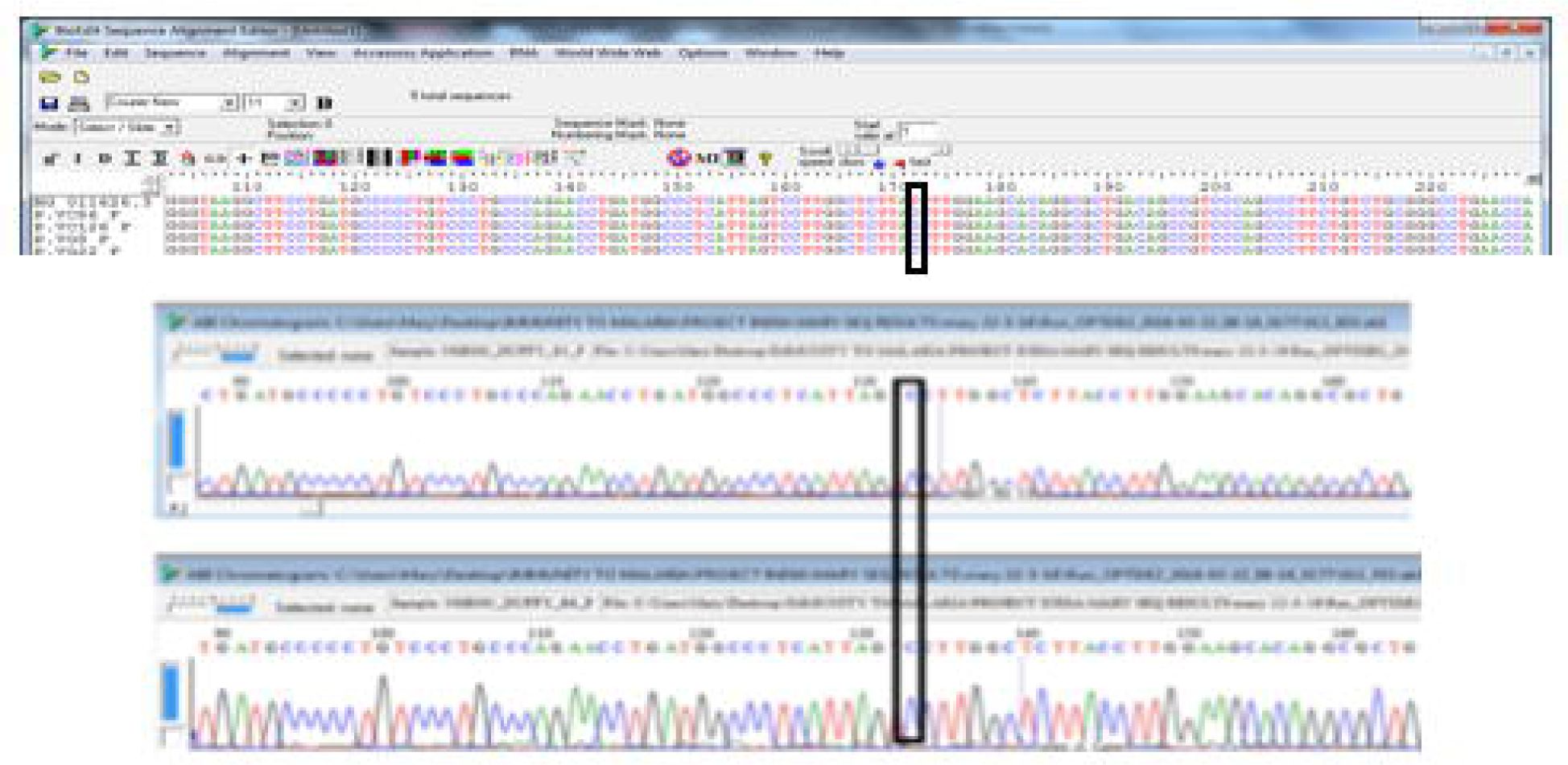
Gel documentation of various *Plasmodium* species. A-: First well – DNA base pair ladder, well 2: NC-negative control template (distill water), well 3-PC-P. *falciparum* positive control, well 4-9-isolates of *P. falciparum*, well 10-DNA base pair ladder, wells 11 and 12-negative and positive controls of *P. vivax*, wells 13-16-*P. vivax* samples, well 17-DNA base pair ladder, 18 and 19-negative and positive controls of *P. malariae*, well 20-the only additional *P. malariae* detected. B-: well 1-DNA base pair ladder, wells 2 and 3-negative and positive controls of *P. ovale*, wells 4-7-*P. ovale* isolates

Authentication of the PCR results were followed by sequencing (in the forward and reverse direction) the 18S rRNA of all additionally identified species with special focus on *P. vivax*. Sequences were edited and aligned (using BioEdit) with their respective reference strain: *P. vivax* with the Sal-1 strain (accession number U03079.1), *P. falciparum* with the *3D7* strain (accession umber XR_002273095.1), *P. ovale* (accession number L48987.1) and *P. malariae* (accession number NG_011626.30). Surprisingly, all sequences showed perfect homology (100% similarity) with their references as expected (Figur 2-for *P. vivax* and supplementary Figures 1, 2 and 3). This is not unexpected as cases of *P. vivax* have been identified in many countries in sSA (3–5,29,33,39–42) including Nigeria (6), where it was thought to be absent due to the non-expression of the DARC gene on the RBC of majority of the population.Thus, this is adding to the growing evidence of the proposed gradual incursion of *P. vivax* into sSA sub-region (data unpublished).

**Figure 2:**
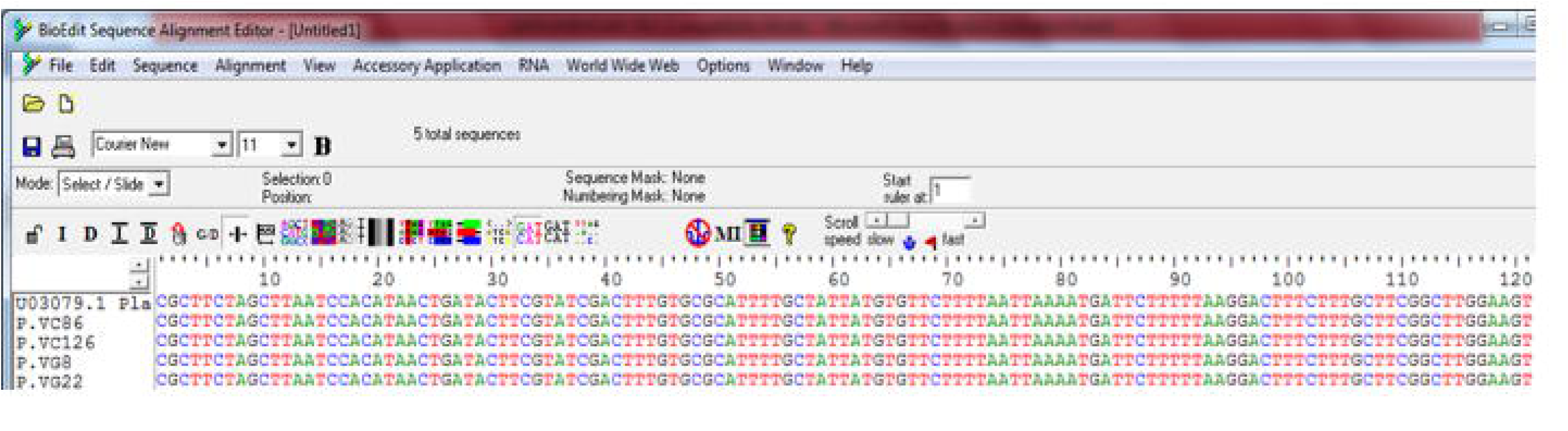
Multiple sequence alignment of *P. vivax* isolates and its Sal -1 reference sequence. In order to discern the Duffy status of those *P. vivax* infected patients, a portion of the DARC gene (precisely the promoter region covering the T33C point mutation) was amplified and sequenced follwing previous protocol (43). Unanticipatedly, all four additional *P. vivax* isolates carried a single cytosine (C) peak at the 33^rd^ nucleotide position upstream (Figure 3). Thus, confirming that none of them expressed the Duffy gene on their RBCs and as such are Duffy negative. All sequences from the current study have been submitted to Genbank (……).

**Figure 3:**
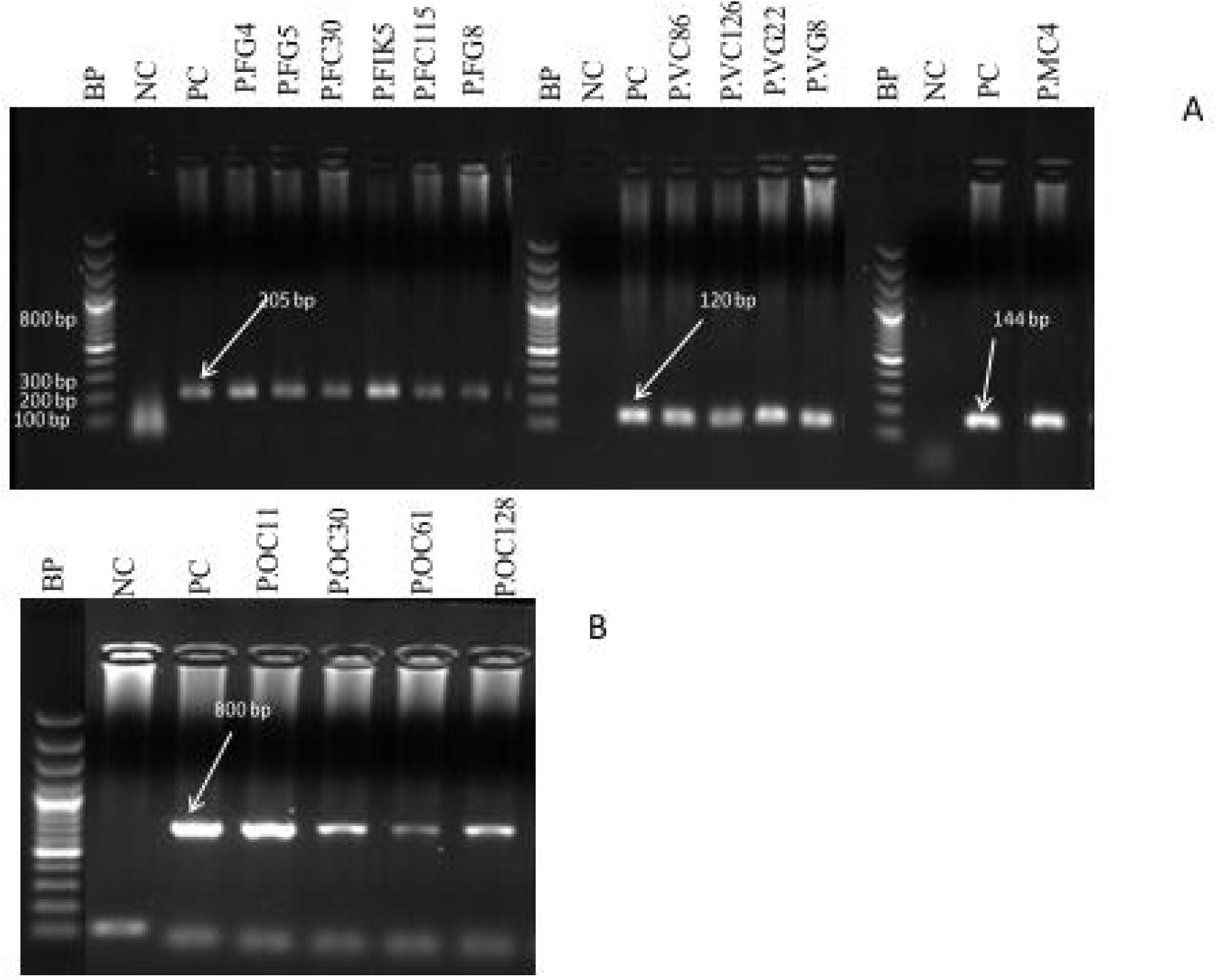
Multiple sequence alignment of the Duffy gene of the vivax samples displaying the –T33C nucleotide change which validates their Duffy negative status. The identification of more *P. vivax* isolates among these Duffy-negative individuals from Nigeria, substantiate the expanding body of evidence of the ability of *P. vivax* to infect RBCs that do not express the DARC gene. Although a very recent findings point to another receptor on the reticulocyte-transferrin receptor 1 as a specific *P. vivax* receptor (44), which is being proposed to be an alternate route of entry into the RBCs, this however is subject to further verification. This is one of the proposition being made to support the observation of *P. vivax* in sSA (45). Another hypothesis which relies on the first conjecture (assuming it is agreed that *P. vivax* at least possesses alternative invasion pathway), is that the Duffy positive carriers in northern part of Africa and the Afro-Asiatic populations of Sudan, Somalia (20,46) and Ethiopia (47,48) serve as reservoir to effect transmission to Duffy negative individuals in those areas as well as other countries (in sSA) through migration. Albeit, this particular hypothesis at play here is yet to be determined. One thing is clear here however, that the true epidemiological situation of *P. vivax* in sSA in particular and Africa in general is yet to be ascertained. Therefore, such genetic epidemiological studies should be conducted in other areas (for example this study covers only two states out of the thirty-six in Nigeria) of the country as with other sSA countries. This will aid in putting in place appropriate control strategies to combat the menace of malaria infection in this most affected population (Africa) and also prevent further spread of *P. vivax* in Africa.

## Data Availability

All data except the sequence stated in this manuscript are embedded in the manuscript.
Sequences would be submitted to the NCBI following manuscript approval.

## Acknowledgements

We thank all the study participants for consenting to donate blood samples for this study. OMA acknowledges the Department of Science and Technology Government, and Ministry of External Affairs (MEA), Government of India (GoI), and the Federation of Indian Chambers of Commerce and Industry (FICCI), for awarding the Visiting Fellowship under the CV Raman Fellowship scheme. Thanks are also due to the Rector of the Université Cheikh Anta Diop de Dakar, Senegal and Director of ICMR -National Institute of Research in Tribal Health (NIRTH) for providing instrumental facilities. We thank Mrs. Ayanlere, Central Hospital, Nigeria, and Mr. Agbayewa, Ikate Primary Health Care, Nigeria for helping in the collection of samples and Mr. Sri Krishna and Ms. Priyanka Patel, ICMR-NIRTH, India for assistance in the laboratory.

## Supporting Information

**S1 Table. PCR cycling conditions for all *Plasmodium* species**

**S2 Table. Primer sequence and cycling conditions for the Duffy negative antigen**

## Notes

### Competing Interest Statement

The authors have declared no competing interest.

### Author Declarations

All relevant ethical guidelines have been followed and any necessary IRB and/or ethics committee approvals have been obtained.

Any clinical trials involved have been registered with an ICMJE-approved registry such as ClinicalTrials.gov and the trial ID is included in the manuscript.

